# Low knowledge about hepatitis B prevention among pregnant women in the Democratic Republic of the Congo

**DOI:** 10.1101/2022.04.12.22273794

**Authors:** Sahal Thahir, Samantha E. Tulenko, Patrick Ngimbi, Sarah Ntambua, Jolie Matondo, Kashamuka Mwandagalirwa, Martine Tabala, Didine Kaba, Marcel Yotebieng, Jonathan B. Parr, Peyton Thompson

**Affiliations:** Department of Pediatrics, University of North Carolina at Chapel Hill; Department of Epidemiology, UNC Gillings School of Global Public Health; Kinshasa School of Public Health; Division of General Internal Medicine, Department of Medicine, Albert Einstein College of Medicine; Department of Medicine, Division of Infectious Diseases, University of North Carolina at Chapel Hill; Department of Pediatrics, Division of Infectious Diseases, University of North Carolina at Chapel Hill

## Abstract

Infants infected perinatally with hepatitis B (HBV) are at the highest risk of developing chronic hepatitis and associated sequelae. Prevention of mother-to-child transmission (PMTCT) of HBV requires improved screening and awareness of the disease. This study evaluated existing HBV knowledge among pregnant mothers (n = 280) enrolled in two HBV screening studies in two urban maternity centers in Kinshasa, Democratic Republic of the Congo. All mothers responded to three knowledge questions upon study enrollment. Baseline levels of knowledge related to HBV transmission, treatment, and symptoms were low across all participants; 60% of participants responded “I don’t know” to all questions. HBV-positive women who participated in both studies (n= 46) were asked the same questions during both studies and showed improved knowledge after screening and treatment, despite no formal educational component in either study. These findings highlight the need for intensified education initiatives in highly endemic areas to improve PMTCT efforts.

## Introduction

Hepatitis B virus (HBV) is one of the most common causes of chronic hepatitis in the world, with highest prevalence in sub-Saharan Africa (SSA) and the Western Pacific region.^1^ Despite an effective vaccine, an estimated 1.5 million new infections occurred globally in 2019, with almost 1 million new infections occurring in Africa.^1^ Most chronic infections in SSA result from HBV transmission that occurs before adolescence, either from mother-to-child transmission (MTCT) or horizontal transmission among children.^2^ Prevention of MTCT (PMTCT) is critical to addressing the HBV epidemic,^3^ and increased knowledge about HBV could improve women’s participation in prenatal screening and PMTCT measures.

Chronic HBV is highly endemic in the Democratic Republic of the Congo (DRC).^4^ Currently, the DRC is evaluating potential HBV screening, birth-dose vaccination, and treatment programs for pregnant women in the DRC. Here, we assess HBV knowledge and perceptions among pregnant mothers in Kinshasa, DRC in preparation for a forthcoming roll-out of a national hepatitis prevention plan.

## Methods

In this study, we performed a secondary analysis of data collected during two studies of HBV prevention of maternal-to-child transmission (PMTCT) in Kinshasa, DRC: The Arresting Vertical Transmission of HBV (AVERT) study^5^ and the ongoing Birth Dose Immunogenicity (BDI) study (**Figure 1**). Both studies were conducted in two large, private, not-for-profit facilities: Kingasani and Binza maternity centers. The AVERT study was developed to demonstrate the feasibility of HBV PMTCT through testing/treating pregnant women and birth-dose vaccination for exposed infants. The aim of the BDI study (unpublished) is to determine the immunogenicity of an added HBV birth-dose vaccination to the infant immunization schedule in HBV-exposed and HBV-unexposed infants. Mothers in the AVERT study were HBV-positive women who enrolled during prenatal visits in 2018-2019. Mothers in the BDI study included previously identified HBV-positive women from AVERT who consented to join BDI as well as newly screened, mostly HBV-negative women who were enrolled when they presented for delivery in 2019 (**Figure 1**). Upon initial study enrollment, participants received group counseling on HBV and PMTCT and underwent point-of-care HBV surface antigen (HBsAg) testing. Participants also completed a standardized questionnaire on demographics and clinical characteristics; this questionnaire included a section on HBV knowledge regarding modes of transmission, signs and symptoms of infection, PMTCT measures, and perceived severity of HBV infection (**Supplementary Exhibit 1**). Mothers who participated in both AVERT and BDI completed the questionnaire at AVERT enrollment and again approximately 9 months later at BDI enrollment. Institutional review board approval was obtained from the University of North Carolina (17-2090 [AVERT] and 18-2793 [BDI]) and from Kinshasa School of Public Health (ESP/CE/021/2019 and ESP/CE/062/2019, respectively). Both studies were registered at clinicaltrials.gov (NCT03567382 and NCT03897946, respectively). Written informed consent was obtained from all subjects.

**Fig 1.**
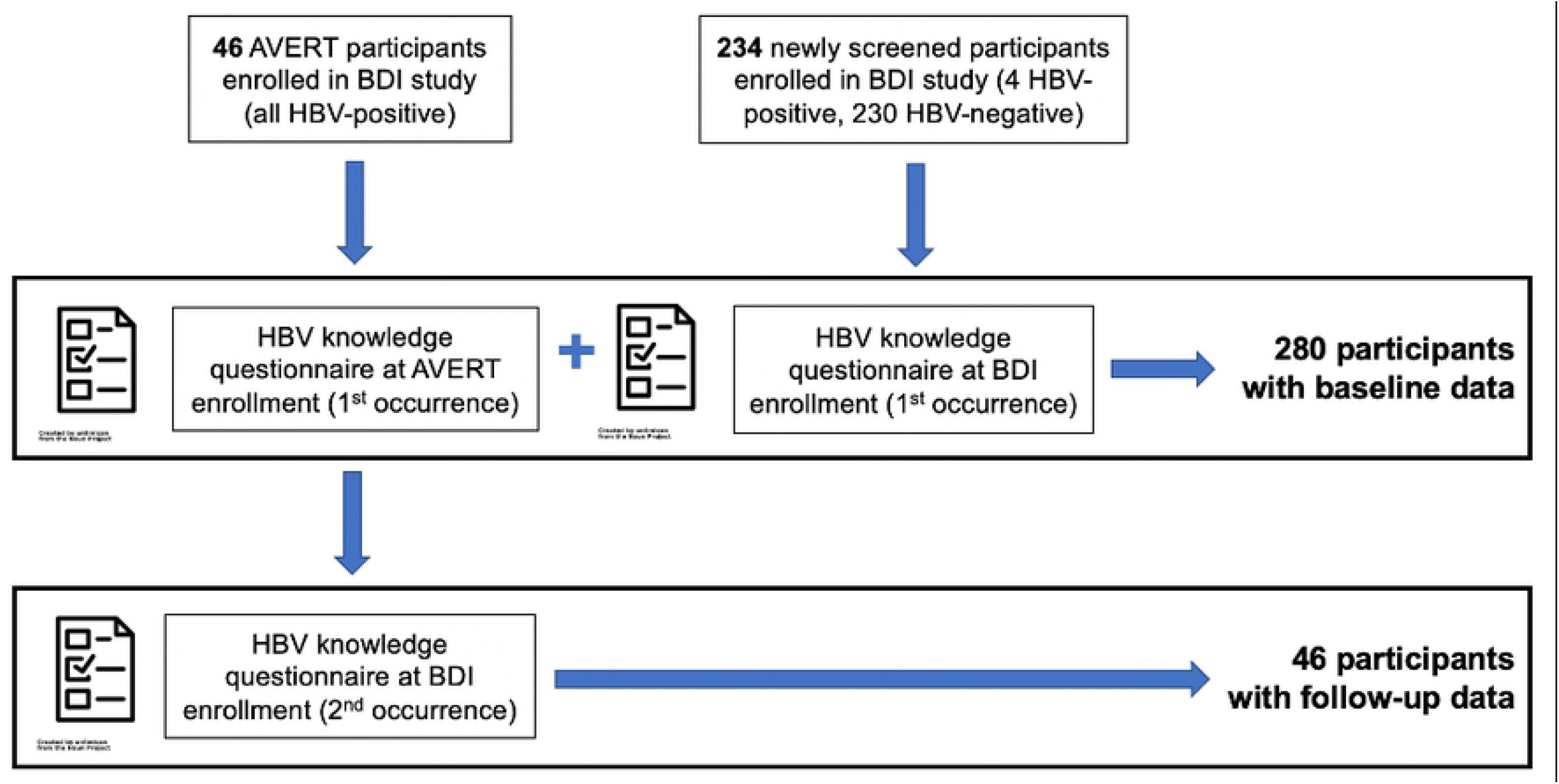
Schematic depicting activities in the AVERT and BDI studies, including the point at which enrollment questionnaires were conducted.

HBV knowledge scores were calculated as the sum of up to 9 correct answers to 3 knowledge questions on the baseline questionnaire (**Supplementary Exhibit 1**). We assessed baseline HBV knowledge as well as demographic characteristics using descriptive statistics. Among AVERT mothers, we compared pre- and post-enrollment HBV knowledge scores using a paired t-test. All analyses were conducted using R (Version 1.4.1717).

## Results

The study population consisted of 280 pregnant women recruited from Binza and Kingasani maternity centers in Kinshasa Province as part of the AVERT and BDI trials. Fifty women (17.9%) were HBV-positive and 230 (82.1%) were HBV-negative. By design, approximately half of the women were recruited from each maternity center. Of the 50 HBV-positive women, 46 (92.0%) were enrolled in the AVERT trial prior to participating in the BDI study and 4 (8.0%) screened positive during BDI enrollment. The subset of women enrolled in both the AVERT and BDI studies completed the questionnaire twice and were assessed for both baseline HBV knowledge and change in HBV knowledge following participation in the AVERT study (**Figure 1**). Responses from the remaining 234 women (230 HBV-negative, 4 HBV-positive) who completed the knowledge questionnaire upon BDI study enrollment were analyzed only for baseline HBV knowledge.

On the baseline knowledge assessment, the average HBV knowledge score among all women was 1.08. Scores ranged from 0 to 6 out of 9; no participants answered all questions correctly. The majority of women (60.0%) had no correct answers, receiving a score of 0 on our 9-point HBV knowledge assessment scale (**Figure 2**). All of these women responded “I don’t know” in response to each HBV question. Within each section of the questionnaire, most women responded that they did not know the correct answer: 233 women (79.6%) did not know the signs and symptoms of HBV, 192 women (68.8%) did not know how a person could get HBV, and 198 women (70.7%) did not know how MTCT of HBV could be prevented (**Table 1**). Women with no HBV knowledge were slightly younger, more likely to be HBV-negative, more likely to attend Binza maternity center, and less likely to have worked in the last 7 days (**Table 2**). The majority (97.5%) of women had attended secondary school or a higher education level.

**Table 1.** Responses to the three questions assessing Hepatitis B knowledge among all enrolled women. 280 total participants responded to each question. Participants could choose multiple answers to each question. Percentages for the individual questions are calculated out of the total number of participants for each question, not total number of answers chosen, so percentages sum up to >100%.

| Select all that apply: | Number (%) of participants who chose this answer |
| --- | --- |
| No symptoms | 3 (1.1%) |
| Jaundice (yellowing of skin) | 42 (15.0%) |
| Abdominal pain | 18 (6.4%) |
| Rash | 9 (3.2%) |
| Don't know | 223 (79.6%) |
| Refuse to answer | 0 (0%) |

| Select all that apply: | Number (%) of participants who chose this answer |
| --- | --- |
| Handshakes | 2 (0.7%) |
| Contact with infected person's blood | 63 (22.5%) |
| Sharing dishes | 1 (0.4%) |
| Mother-to-child transmission | 25 (8.9%) |
| Sexual transmission | 48 (17.1%) |
| Breastfeeding | 0 (0%) |
| Don't know | 192 (68.6%) |
| Refuse to answer | 1 (0.4%) |

| Select all that apply: | Number (%) of participants who chose this answer |
| --- | --- |
| Infant vaccination at birth | 42 (15.0%) |

**Table 1.** Responses to the three questions assessing Hepatitis B knowledge among all enrolled women.
|  |  |
| --- | --- |
| Antiviral treatment during pregnancy | 51 (18.2%) |
| Don't know | 198 (70.7%) |
| Refuse to answer | 1 (0.4%) |

**Table 2.** Demographic characteristics of enrolled women in the overall cohort and stratified by HBV knowledge. “No HBV knowledge” is defined as a score of 0 on HBV knowledge assessment and “any HBV knowledge” is a score > 0 on HBV knowledge assessment.

| Characteristic | Overall<br>(n=280) | Any HBV Knowledge<br>(n = 112) | No HBV Knowledge<br>(n = 168) |
| --- | --- | --- | --- |
| Mean age in years (range) | 28.7 (18-45) | 29.5 (18-42) | 28.1 (18-45) |
| Maternal HBV status |  |  |  |
| HBV-positive | 50 (17.9%) | 38 (33.9%) | 12 (7.1%) |
| HBV-negative | 230 (82.1%) | 74 (66.1%) | 156 (92.9%) |
| Maternity center attended |  |  |  |
| Binza | 143 (51.1%) | 17 (15.2%) | 126 (75.0%) |
| Kingasani | 137 (48.9%) | 95 (84.8%) | 42 (25.0%) |
| Gravidity |  |  |  |
| 1-2 pregnancies | 149 (53.2%) | 53 (47.3%) | 96 (57.1%) |
| > 2 pregnancies | 131 (46.8%) | 59 (52.7%) | 72 (42.9%) |
| Marital status |  |  |  |
| Married | 222 (79.3%) | 86 (76.8%) | 136 (81.0%) |
| Unmarried | 58 (20.7%) | 26 (23.2%) | 32 (19.0%) |
| Education |  |  |  |
| No school | 2 (0.7%) | 2 (1.8%) | 0 |
| Primary school | 5 (1.8%) | 2 (1.8%) | 3 (1.8%) |
| Secondary school | 200 (71.4%) | 80 (71.4%) | 120 (71.4%) |
| Higher education | 73 (26.1%) | 28 (25.0%) | 45 (26.8%) |
| Work in the last 7 days |  |  |  |
| No work | 175 (62.5%) | 60 (53.6%) | 115 (68.5%) |
| Any work | 105 (37.5%) | 52 (46.4%) | 53 (31.5%) |
| Wealth quartile |  |  |  |
| Lowest quartile | 70 (25.0%) | 24 (27.4%) | 46 (21.4%) |
| Lower-to-middle | 70 (25.0%) | 22 (28.6%) | 48 (19.6%) |
| Upper-to-middle | 70 (25.0%) | 32(22.6%) | 38 (28.6%) |
| Highest quartile | 70 (25.0%) | 34 (21.4%) | 36 (30.4%) |

**Fig 2.**
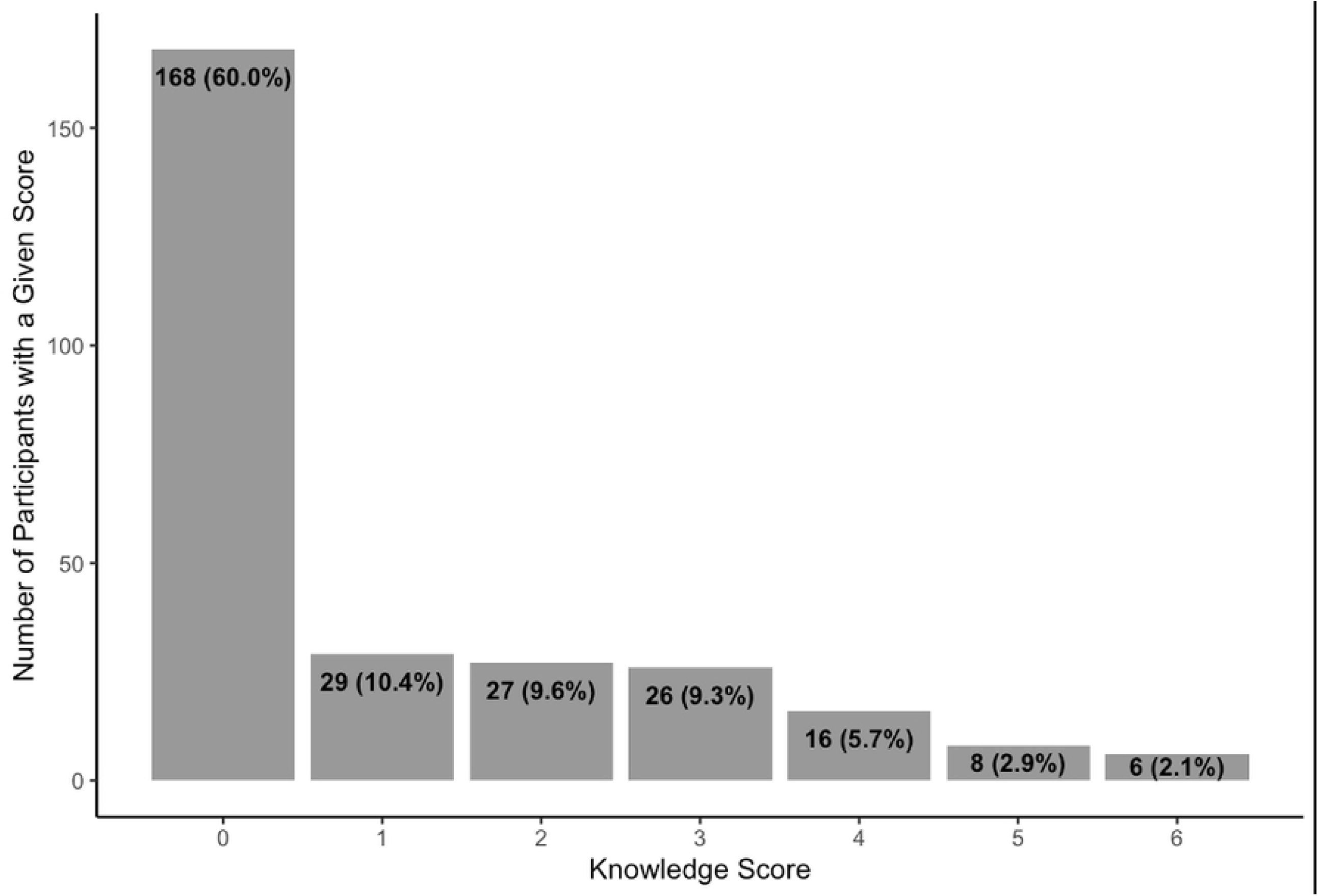
Distribution of HBV knowledge scores at baseline. Score reflects the number of correct responses to 9 statements about HBV symptoms, transmission, and prevention. The average knowledge score was 1.06.

In response to the question, “In your opinion, how serious a disease is Hepatitis B?”, most women (70.7%) selected “very serious” and 20.7% selected “I don’t know” (**Figure 3**). A greater proportion of women with any HBV knowledge considered HBV “very serious” compared with women with no HBV knowledge (77.7% vs. 66.1%). More women with no HBV knowledge responded that they “didn’t know” about HBV severity (28.6% vs. 8.9%).

**Fig 3.**
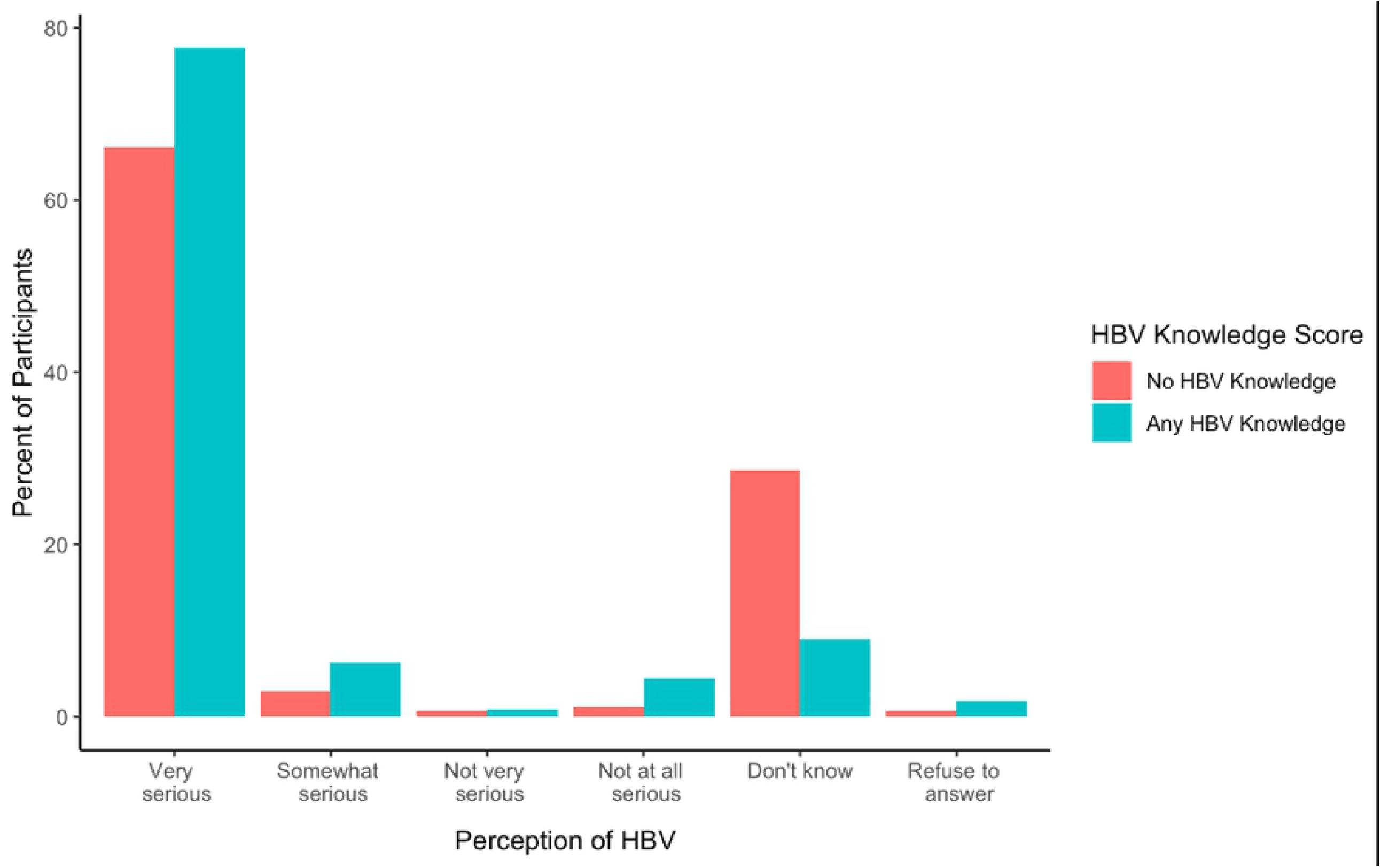
Participants responses to the question “in your opinion, how serious a disease is hepatitis B?” among women with no HBV knowledge (score of 0 on HBV knowledge assessment) and women with any HBV knowledge (score > 0 on HBV knowledge assessment).

Among the 46 women who participated in both studies, the average knowledge score at follow-up was significantly higher than the average score at baseline (3.7 vs. 2.8, p = 0.004) (**Figure 4**). Three answer choices in particular had a higher proportion of correct responses at follow-up compared to baseline: (1) recognizing jaundice as a symptom of HBV (52.2% correctly reported at baseline vs. 91.3% at follow-up), (2) recognizing that a person can contract HBV through contact with blood from an infected person (54.3% vs. 89.1%), and (3) recognizing that MTCT of HBV can be prevented by infant vaccination at birth (15.2% vs. 76.1%). At follow-up, fewer women responded “I don’t know” to all questions (8 women at baseline [17.4%] vs. 1 woman at follow-up [2.2%]).

**Fig 4.**
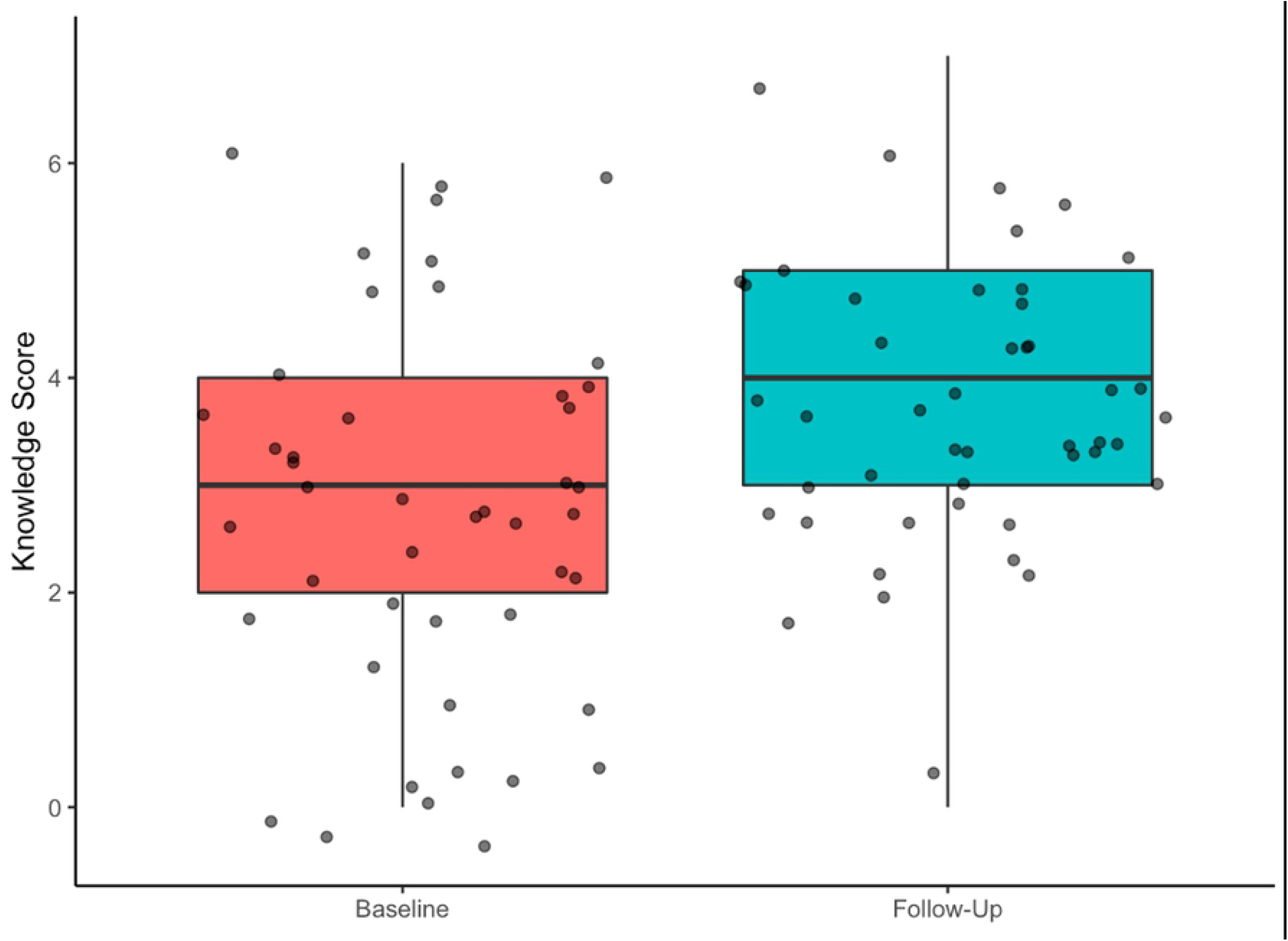
Distribution of knowledge scores at baseline and follow-up among 46 HBV-positive women who participated in both studies. Average score at baseline was 2.8 (range 0-6) and average score at follow-up was 3.7 (range 0-7) (p = 0.004).

## Conclusions

In this analysis of data from two HBV PMTCT studies in Kinshasa, DRC, HBV knowledge was low among pregnant women, with nearly two-thirds of women responding “I don’t know” to all questions asked. This finding is consistent with previous studies of pregnant populations in sub-Saharan Africa, in which 50% to 76% of participants had low HBV knowledge.^6–11^ In a recent study from a rural area of DRC, authors reported even less baseline HBV knowledge: 93% of study participants had no knowledge of HBV.^12^ Mudji et al. attributed this low knowledge in part to less health education in more rural areas. Studies in Cameroon, Ethiopia, Kenya, and Ghana found that higher levels of HBV knowledge were associated with increased level of education attained.^8,9,11,13^ In particular, Gebrechokos et al. noted that factors such as education level and higher monthly income were associated with adequate HBV knowledge, likely due to improved access to media and technologies allowing for access to HBV-related information^11^. However, in our study, low education did not appear to explain lack of HBV knowledge, as 97.5% of participants had attended at least secondary school (**Table 1**). Instead, low HBV knowledge might be due to a lack of public education regarding HBV in the DRC, highlighting an important gap in public health messaging.

Encouragingly, we observed improvement in HBV knowledge over time among HBV-positive mothers who participated in the AVERT trial.^13^ These women received brief, informal HBV education upon enrollment, and participation in study activities might also have passively increased awareness of HBV. Alternatively, this knowledge increase may have been due to frequent interactions with healthcare workers during the treatment period, which has been shown to improve healthcare worker trust and communication.^14–16^ Participation in the study may have increased these women’s confidence and comfort in interacting with study staff and in responding to the questionnaire. Of note, HBV-positive women had higher average knowledge scores than HBV-negative women at baseline. Further research is needed to understand the mechanism for this observed increase in knowledge to inform future educational interventions.

While these findings provide pertinent insight into HBV knowledge levels among pregnant women in Kinshasa, this study is limited by its retrospective design and use of existing data from study questionnaires that were not originally designed to comprehensively assess HBV knowledge and perceptions. Additionally, study participants were recruited from large maternity centers in urban Kinshasa, and findings may not be generalizable to other areas of the DRC where access to healthcare and public health education is generally more limited. However, given the low knowledge scores among this relatively well-educated cohort, it is reasonable to infer that HBV knowledge may be even lower in other settings in the DRC.

The low baseline HBV knowledge scores and their improvement over time observed in this study suggest that HBV education efforts for pregnant women should be prioritized as part of HBV elimination efforts. The improvement in HBV knowledge observed over the course of the AVERT study suggests that one strategy might be to integrate HBV testing and treatment into routine prenatal care alongside a rigorous educational initiative. Future studies that pilot HBV educational interventions for pregnant women are urgently needed in sub-Saharan Africa.

## Data Availability

Upon manuscript acceptance, data will be made publicly available through the Carolina Digital Repository (accessible at https://cdr.lib.unc.edu/catalog?locale=en)

## Acknowledgements

We thank all of the women who participated in this study, the staff at the Binza and Kingasani maternity clinics, and provincial and national health authorities. We would like to thank Dr. Noro Ravelomanana, Dr. Malongo Fathy, and Dr. Bienvenu Kawende for their contributions to this study.

We are grateful for the support from the administrative staff at the Kinshasa School of Public Health and at the University of North Carolina. We grieve the loss of Dr. Steven Meshnick, whose vision and mentorship were critical to the success of this study.

